# Vertebral Bone Infarction: A Key Clue in Identifying Spinal Cord Strokes

**DOI:** 10.64898/2025.12.03.25341600

**Authors:** Paula Barreras, David Acero-Garces, Lydia Gregg, Hao Wen Chen, Karissa Arthur, Philippe Gailloud, Doris Lin, Carlos A. Pardo

**Affiliations:** Department of Neurology, Johns Hopkins University School of Medicine, Baltimore, MD, USA; Department of Neurology, Cedars Sinai Medical Center, Los Angeles, CA, USA; Department of Art as Applied to Medicine, Johns Hopkins University School of Medicine, Baltimore, MD, USA; Clinical Neurology, Lewis Katz School of Medicine at Temple University, Philadelphia, PA, USA; Department of Radiology, Johns Hopkins University School of Medicine, Baltimore, MD, USA; Department of Pathology, Johns Hopkins University School of Medicine, Baltimore, MD, US

**Author notes:** **Corresponding author:** Carlos A. Pardo, MD, Department of Neurology, Johns Hopkins School of Medicine 627 Pathology Building, 600 North Wolfe Street, Baltimore, Maryland, 21287.

**Keywords:** spinal cord ischemia, vertebral body infarction, vascular myelopathy, vascular supply

## Abstract

**Background:** Spinal cord stroke (SCS) is rare and frequently misdiagnosed. This study describes the clinical and radiological characteristics of vertebral body infarction (VBI) associated with SCS.

**Methods:** To assess VBI in patients with SCS, we retrospectively examined clinical data and magnetic resonance imaging from patients diagnosed at a specialized referral center. VBI was defined as a T2-weighted hyperintensity within a vertebral body near ischemic cord lesions. Additional features, such as gadolinium enhancement, lesion distribution, and the exclusion of other explanations, were also analyzed. Each vertebra was evaluated for its level, pattern of signal changes, and enhancement, then correlated with the lesion distribution and related arterial spinal supply.

**Results:** Among 126 patients with SCS, 15 (12%) showed MRI evidence of VBI. Thirteen patients (87%) had multiple vertebrae affected, with 80% of cases involving two adjacent vertebral bodies. Of the 35 vertebrae displaying signs of VBI, 20 (57%) were thoracic, 9 (25%) were cervical, and 6 (17%) were lumbar. In 80% of SCS cases, VBI MRI abnormalities were found at the same or lower vertebral levels corresponding to the cord lesions; these were typically hyperintense on T2-weighted and STIR sequences, with 92% showing contrast enhancement. The location of the ischemic lesions in the vertebral bodies was anterolateral (47%), posteramedial (23%), hemivertebral (17%), or posterolateral (9%), with two vertebrae showing diffuse involvement. The spinal vascular territories included the artery of Adamkiewicz (n=9, 60%), the upper artery of the cervical enlargement (n=6, 40%), the artery of von Haller (n=5, 33%), and the lower artery of the cervical enlargement in two cases (13%).

**Conclusion:** The co-occurrence of VBI and SCS is attributable to their shared arterial supply, as the MRI findings in VBI generally mirror the spinal vascular anatomy. VBI serves as a key indicator of vascular injury and an important diagnostic marker for SCS.

## INTRODUCTION

Spinal cord stroke (SCS) is a rare condition compared to its brain counterpart. It can occur due to traumatic injury to the spinal cord blood supply or spontaneously as a result of various risk factors that affect the arterial blood supply to the spinal cord in the cervical, thoracic, or lumbar regions of the spine. Spontaneous SCS is often misdiagnosed as “transverse myelitis,” with up to 20% of cases initially diagnosed as inflammatory myelopathy later identified as vascular myelopathy. ^1,2^ Misdiagnosis frequently results from a lack of awareness of specific MRI patterns associated with SCS and the clinical overlap among different myelopathies.

Misunderstandings of the vascular anatomy of the spinal cord can further complicate diagnosis, leading to low suspicion when lesions do not match the typical distribution seen in the “classic” syndromes associated with the vertebral and Adamkiewicz arteries or the anterior or posterior spinal arteries. These classic syndromes, which involve the anterior two-thirds and posterior third of the spinal cord, are rarely documented in clinical practice. Vertebral body infarction (VBI), reported in 7% to 35% of patients with SCS ^3,4^, serves as an indirect marker of SCS. Although there are no unified criteria in the literature to define VBI on MRI, and no single imaging finding can be used to diagnose it, previous reports have outlined some of the imaging features of SCS. ^5,6^ VBI may be the only abnormality identified on the initial spinal MRI during the acute stage ^5^, but it can also appear hours to weeks after SCS begins. ^6,7^ The coexistence of VBI and SCS occurs because of the shared arterial supply between the spinal cord and vertebral column (**Figure 1**). While the vertebral artery supplies most of the blood to the cervical spine vertebrae, the thoracic and lumbar vertebrae receive blood from intersegmental arteries (ISAs) that branch off the aorta and run along the anterolateral aspect of the vertebral bodies.

**Figure 1.**
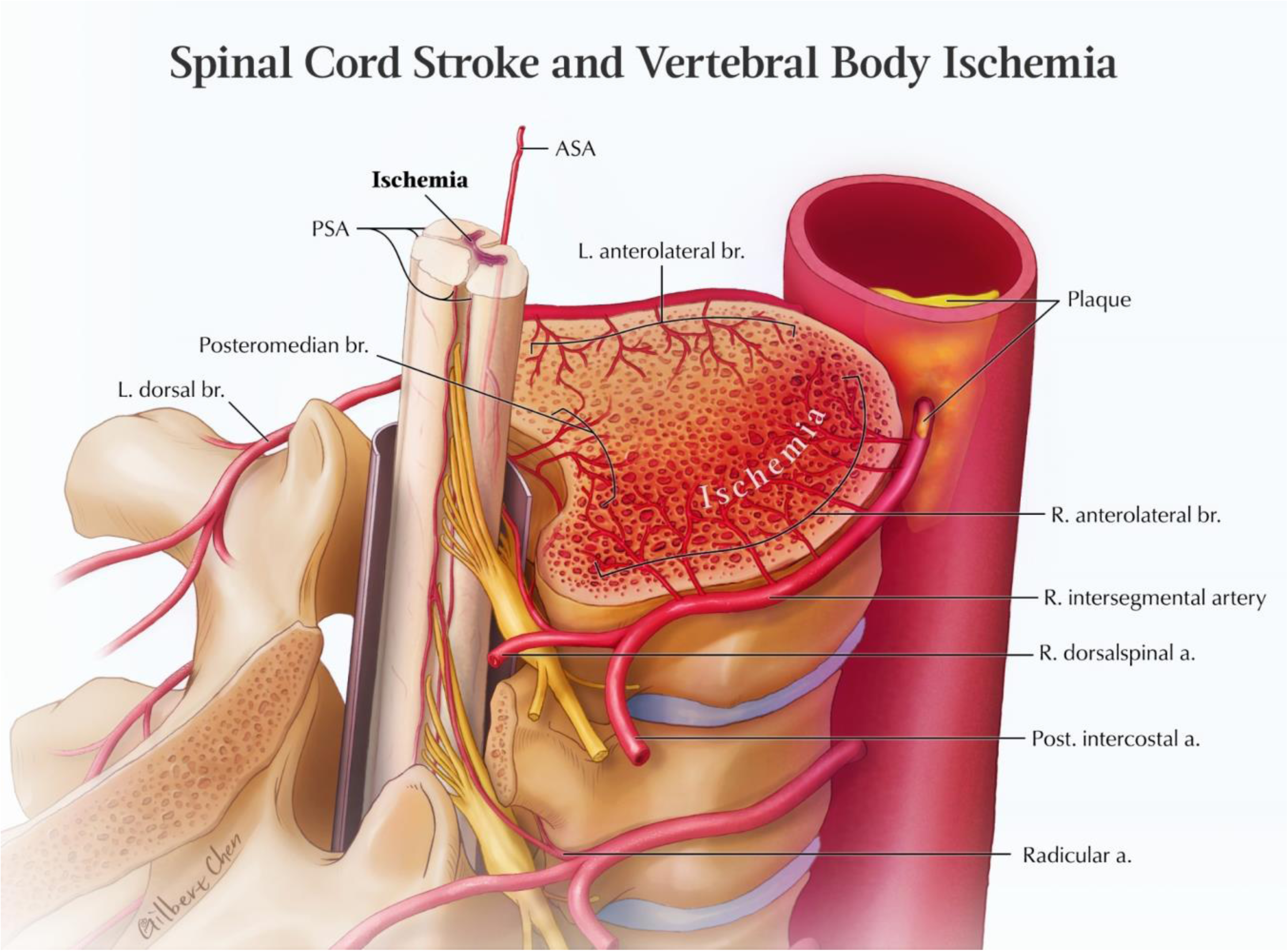
Vascular supply to a thoracic vertebral body and spinal cord. This schematic illustration depicts the vascular supply to a vertebral body and spinal cord. The intersegmental arteries (ISA), which arise from the aorta, branch out to supply the anterolateral aspects of the vertebral body (R. and L. anterolateral br.). ISAs exhibit variations, including bilateral and unilateral trunks. Bilateral trunks, which give rise to both left and right ISAs for a single vertebra, are more commonly found in the lower lumbar region. In contrast, unilateral trunks supply the ipsilateral ISAs of two or more adjacent vertebrae, predominantly in the upper thoracic region. Each ISA divides into a lateral and a dorsospinal branch. The lateral branch provides blood supply to the thoracic and abdominal walls (Post. Intercostal a.), while the dorsospinal branch gives the dorsal branch (L. dorsal br.), primarily supplying the posterior vertebral elements, including the outer aspect of the lamina and the spinous process, and the paraspinal musculature.^8^ A spinal branch originates directly from the ISA or its dorsospinal branch. This spinal branch crosses the neural foramen, dividing into two main branches: the radicular (or radiculomedullary) artery (Radicular a.), which follows the nerve root, and the retrocorporeal artery, which supplies the posterior portion of the vertebral body through the posteromedian arteries (Posteromedian br.). Both anterior and posterior radiculomedullary arteries contribute to the formation of the anterior (ASA) and posterior spinal arteries (PSA), which are the primary blood supplies to the spinal cord. Occlusion of the proximal ISA, whether caused by atherosclerosis (plaque) or an extruded anterolateral or posterior disc, can result in ischemia affecting both the vertebral body and the spinal cord at the same level. The angioarchitecture of the cervical spine differs (not illustrated here), as cervical ISAs remodel into paired vertebral arteries. The anterior aspect of the cervical vertebrae is supplied by branches of the ventral vertebral artery, remnants of the ISA stems, as well as subclavian branches (in the caudal region) and external carotid branches (in the cranial region); these branches typically do not contribute to the spinal cord vascularization. The posterior aspects of the first five or six cervical vertebrae receive blood supply from retrocorporeal arteries arising from the vertebral arteries, while the lower cervical vertebrae (typically C7, sometimes C6) are supplied by subclavian branches via the supreme intercostal artery. Abbreviations: ASA: anterior spinal artery; PSA: posterior spinal artery; R: right; L: left; br.: branch; a: artery.

These networks supply the anterolateral portions of the vertebrae, whereas other arteries supply the posterior portions. Pathologies affecting these blood supply systems can cause ischemic injury to both the vertebrae and the spinal cord, which can be seen on neuroimaging. Thus, detecting VBI can be a key indicator of vascular pathologies when evaluating myelopathies and supports the diagnosis of SCS. This study reports the frequency of VBI in a large cohort of patients with spontaneous SCS cases seen at a specialized center. It discusses the relevant vascular anatomy, regional differences, and their significance for diagnosis.

## METHODS

We retrospectively examined clinical data and magnetic resonance imaging (MRI) from patients diagnosed with SCS who were referred to a specialized center from 2010 to 2022.

The diagnosis of SCS was based on clinical and neuroimaging criteria ^1^, requiring a clinical syndrome consistent with myelopathy and a spinal cord abnormality documented by MRI in a known arterial territory or watershed area. SCS diagnoses were classified as *definite* when a vascular lesion, such as an occluded artery, was documented angiographically, as *probable* when diffusion restriction was observed on diffusion-weighted imaging (DWI) sequences, or a stroke mechanism was identified, or as *possible* when the clinical profile was consistent with SCS but no stroke mechanism was identified, and other etiologies were ruled out. Diagnoses were determined by a neurologist with clinical expertise in SCS and other myelopathies [CP]. Patients were included in the study if their MRI and clinical records were available for review. Clinical and neuroimaging features were recorded in a REDCap database. Clinical data comprised patient demographics, temporal profile of symptom evolution, presenting symptoms, clinical examination findings, and identified mechanisms for SCS. All available MRIs obtained less than one month after symptoms onset were reviewed to identify the presence of VBI; if more than one MRI was obtained after one month of symptoms onset, at least one was reviewed. All MRIs were obtained for clinical care. Classification of MRI findings, including the presence and features of SCS and VBI, was achieved by consensus among a neuroradiologist [DL] and two neurologists [CP and PB]. Spinal cord lesions were categorized as longitudinally extensive if they spanned three or more vertebral segments, and either as monofocal or multifocal if there were single or multiple lesions, respectively. The arterial territories supplying the spinal cord were classified depending on the vertebral level of the lesion and categorized based on anatomical location as the territory of the vertebral artery/superior artery of the cervical enlargement (C1-C6), inferior artery of the cervical enlargement (C7-T2), artery of von Haller (T3-T5), or Adamkiewicz artery (T6-L1). ^8^ VBI was defined as a hyperintensity on T2-weighted (T2W) and Short Tau Inversion Recovery (STIR) MRI sequence in a vertebral body at or near the level of the cord lesion; supporting findings such as matching enhancement in T1-weighted with Gadolinium sequences, a wedge-shaped, irregular (also described as “geographic”) signal change, and a lesion topography matching a territory of osseous blood supply. Spinal cord and vertebral bone diffusion were also assessed using diffusion-weighted imaging (DWI) when available. Special attention was paid to exclude imaging findings consistent with other vertebral bone lesions that may appear similar on MRI, such as degenerative disease or hemangiomas. For each VBI, the vertebral level, presence of Gadolinium enhancement, and infarction distribution in each vertebra, including the vertebral body and processes, were recorded. To correlate the imaging findings with the anatomical basis, the VBI was assigned to a territory of the osseous arterial supply (Figure 1) based on the distribution of signal intensity changes ^9,10^. The patterns of vertebral body involvement were defined as: 1) anterolateral, when the lesion affected the anterior two thirds or the lateral aspect of the vertebral bone; 2) posterolateral, when the lesion affected the posterior third of the lateral aspect of the vertebral body; 3) posteromedian, when the lesion affected the median aspect of the posterior third of the vertebral body, usually in a wedge shape; 4) hemivertebral, when there was simultaneous anterior and posterior involvement unilaterally; and 5) diffuse, when the signal change was present throughout the vertebral body.

For the descriptive analysis, categorical data were presented using absolute and relative frequencies, and numerical variables were summarized using the median and interquartile ranges. All analyses were performed in R 4.4.2.

### Study protocol approvals and patient consents

This study was conducted with approval from the Johns Hopkins Medical Institutions Institutional Review Board. Patient consent was waived.

## RESULTS

### Clinical features

One hundred twenty-six patients met the criteria for clinical and neuroimaging diagnosis of SCS. Of these, fifteen (12%) were diagnosed with VBI. The selection and diagnosis flow for analysis is outlined in the Supplemental Figure 1. Comparing SCS cases with or without VBI showed no significant differences in age, gender, race, certainty, temporal profile, lesion extent, or involved vascular territory (Table 1). The median age was 55 years (IQR 44-66). Most patients were women (n=11, 73%) and white (n=14, 93%). The diagnostic certainty of SCS among patients with VBI was as follows: definite in 3 cases (20%), probable in 10 cases (67%), and possible in 2 cases (13%). The most common presenting symptoms included hyperacute or acute onset of limb weakness (n=14, 93%) and numbness (n=14, 93%), followed by bowel and/or bladder dysfunction (n=12, 80%), paresthesias (n=11, 73%), and acute back pain (n=10, 67%). Nine patients (60%) presented with lower motor neuron syndrome characterized by flaccid muscle tone and reduced reflexes. The onset of symptoms was hyperacute (within 6 hours from onset to nadir of neurological dysfunction) in 11 cases (73%), acute (6 to 48 hours from onset to nadir) in 3 (20%), and subacute with a stuttering course in 1 case. The clinical and imaging characteristics of each of the patients with VBI included in the analysis are detailed in Supplemental Table 1. Thirteen patients (87%) received the initial diagnosis of “transverse myelitis,” and most received empirical treatments including intravenous methylprednisolone (n=12), plasma exchange (n=3), and intravenous immunoglobulin (n=1), with little to no improvement. Mechanisms of SCS determined retrospectively included atherosclerotic disease (n=5, 33%), arterial dissection (n=2), hypoperfusion due to severe hypotension (n=2), hypercoagulability (n=2), and mechanical compression by an osteophyte (n=1). The underlying cause remained unknown in 3 cases.

**Table 1.** Clinical, demographics and imaging findings in spinal cord strokes with and without vertebral bone infarct.

|  | **Vertebral bone infarction**<br>**n= 15** | **No vertebral bone infarction**<br>**n= 111** | **p-value** | **Test** |
| --- | --- | --- | --- | --- |
| Age, median (IQR) | 55 (44-66) | 53 (29-60) | 0.2179 | * |
| Female, n (%) | 11 (73) | 62 (56) | 0.2684 | ** |
| White race, n (%) | 14 (93) | 77 (69) | 0.0657 | ** |
| **Diagnostic certainty n (%)** |  |  | 0.5101 | ** |
| Definite | 3 (20) | 17 (15) |  |  |
| Probable |  | 64 (58) |  |  |
| Possible |  | 30 (27) |  |  |
| **Temporal evolution to nadir n (%)** |  |  | 0.2255 | ** |
| Hyperacute | 11 (73) | 89 (80) |  |  |
| Acute | 3 (20) | 21 (19) |  |  |
| Subacute | 1 (7) | 1 (1) |  |  |
| **Imaging findings** |  |  |  |  |
| Lesion length, median (Q1-Q3) | 4 (3-7) | 4 (2-6) | 0.2826 | * |
| **Vascular territory, n (%)** |  |  |  |  |
| Vertebral artery | 6 (40) | 50 (45) | 0.7872 | ** |
| Inferior artery of cervical enlargement | 3 (20) | 47 (42) | 0.1582 | ** |
| Artery of von Haller | 8 (53) | 32 (29) | 0.0758 | ** |
| Adamkiewicz artery | 10 (67) | 51 (56) | 0.1716 | ** |

\* Wilcoxon rank sum test
\*\*Fisher's exact test

### Neuroimaging features

A total of 254 MRIs from 126 subjects diagnosed with SCS were reviewed. Among these, 37 MRIs from 15 patients met the criteria for VBI. Details of all the cases diagnosed with VBI are described in Supplemental Table 1. The interval time between symptom onset and the initial MRI varied considerably. VBI was detected across different clinical phases defined as acute, subacute, or chronic, ranging from 5 hours to 30 days after the onset of initial symptoms. The median time from symptom onset to image acquisition was 4 days (IQR 1-7). In 6 cases (40%), the initial MRI did not reveal any spinal cord abnormalities, but lesions became apparent in subsequent MRIs performed four or more days later. Overall, spinal cord lesions were classified as longitudinally extensive in 8 cases (53%) and monofocal in 9 cases (60%). MRI findings showed that lesions were located in the thoracic cord in 11 cases (73%) and the cervical cord in 6 cases (40%). Notably, 2 cases (13%) exhibited simultaneous involvement of both cervical and thoracic cord due to multifocal, longitudinally extensive lesions. The analysis of vascular supply showed that the Adamkiewicz artery (supplying the T6-lumbar cord segments) was the most frequently involved artery, with 9 cases (60%), followed by the upper artery of the cervical enlargement in 6 cases (40%), the artery of von Haller (T3-T5) in 5 cases (33%) and the inferior artery of the cervical enlargement (C7-T2) in 2 cases (13%). Multiple arterial territories were involved in three cases. DWI sequences were obtained in only 4 cases, all of which showed diffusion restriction of the spinal cord, but none of the vertebral bone (**Figure 2**). Gadolinium-enhanced MRI lesions were observed in 7 cases (47%), with 3 of these cases also showing enhancement of the anterior cauda equina nerve roots during the subacute period.

**Figure 2.**
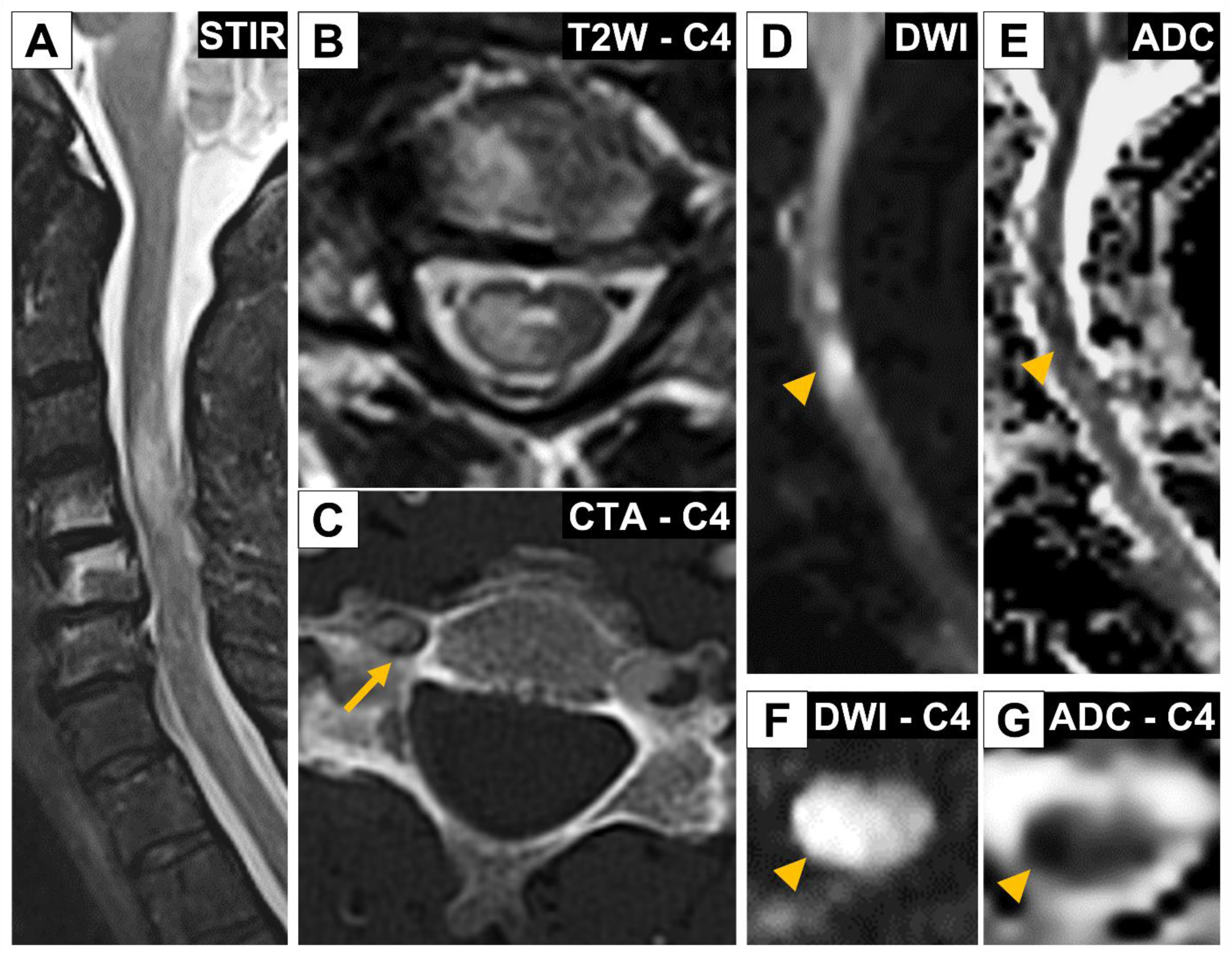
Vertebral bone infarction in the cervical spine. A vertebral bone infarct caused by a right vertebral dissection, initially misdiagnosed as “transverse myelitis” (Case 1). A) Sagittal view of the cervical spine (STIR sequence) reveals a focal area of increased signal intensity in the spinal cord affecting segments C3 to C5. Additionally, increased signal intensities are observed in the inferior aspect of the C4 vertebral body and the superior aspect of C5. B) Axial T2-weighted view at C4 shows a focal area of increased signal intensity within the right hemivertebral region and the right aspect of the cervical spinal cord, consistent with ischemic changes in both the vertebral bone and the spinal cord. C) An axial view of a CT angiogram at C4 reveals a right vertebral artery dissection (arrow). D) Sagittal diffusion-weighted imaging (DWI) of the cervical spinal cord demonstrates focal DWI/ADC signal abnormalities consistent with diffusion restriction. E) Representative axial views at C4 illustrate DWI/ADC signal abnormalities. The signal abnormalities observed in images D-G are consistent with a diagnosis of spinal cord stroke.

MRI evidence of VBI was seen in 35 vertebrae of the 15 patients studied: 9 cervical (25%), 20 thoracic (57%), and 6 lumbar (17%). Multiple vertebrae were affected in 13 cases (87%), typically near the adjacent cord ischemic lesions. The VBIs were found at or below the level of the spinal cord lesions in 12 cases (80%) (Supplemental Figure 2). MRI STIR sequences generally provided more precise visualization of lesions than T2-weighted sequences. The lesions exhibited various configurations, from wedge-shaped (Figure 3) to patchy and poorly delineated (Supplemental Figure 2). Among the Gadolinium-enhanced MRIs obtained from 13 patients, 12 (92%) showed focal vertebral bone enhancement, with the area of enhancement typically smaller than the extent of STIR hyperintensity (Figure 4). The patterns of ischemic vertebral body involvement, correlated with regional arterial blood supply, are illustrated in Figure 5. The ischemic bone lesions were anterolateral in 16 vertebrae (46%), posteromedian in 8 vertebrae (23%) (Figures 3 and 4), and posterolateral in 3 vertebrae (9%) (Figure 6 and Supplemental Figure 3); a hemivertebral distribution was seen in 6 (17%) (Figure 2) and diffuse involvement in 2 (6%) vertebrae (Supplemental Figure 4). In five vertebrae, the VBI extended to the ipsilateral pedicle (three with a posterolateral pattern, one with a hemivertebral pattern, and one with an anterolateral pattern); the spinous process was also involved in one of these cases. MRI T1-weighted images with partial hyperintensity suggested a necrotic-hemorrhagic component in two cases (Supplemental Figure 5). The key imaging features of VBI, compared with those of other non-ischemic vertebral pathologies, such as vertebral body hemangiomas, are summarized in Figure 7.

**Figure 3.**
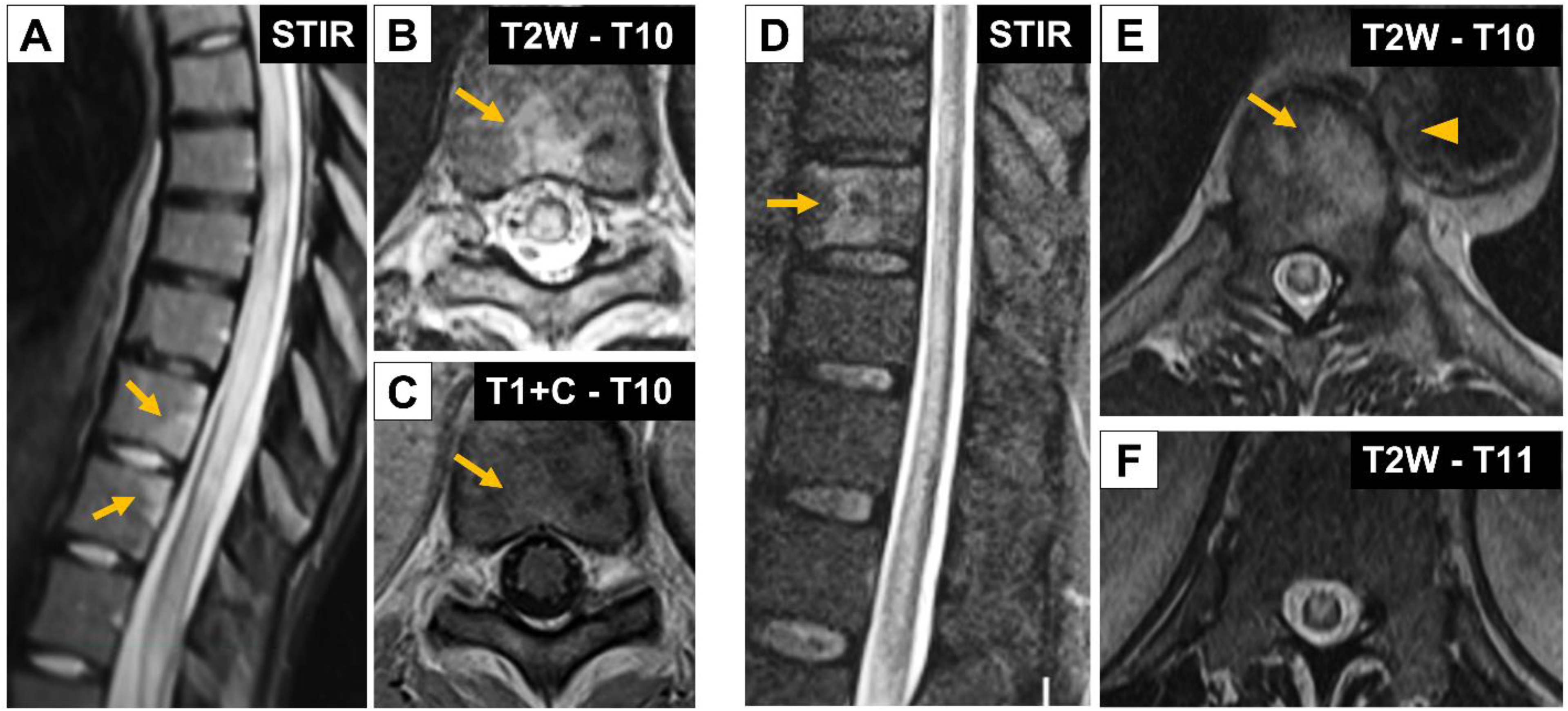
Posteromedian and anterolateral bone infarcts in thoracic spine. A-C) Posteromedian vertebral bone infarction is depicted in sagittal STIR, and axial T2-weighted and Gadolinium-contrasted T1-weighted MR images from Case 15. The images reveal posteromedian signal abnormalities in the T10 and T11 vertebral bodies (A), characterized by a wedge-shaped distribution (arrows) at the lower end of a longitudinally extensive hyperintense spinal cord signal abnormality. The T2-weighted axial image (B) showed the posteromedian distribution of vertebral bone infarction (arrow). In the corresponding T1-weighted axial image (C), mild enhancement is observed following Gadolinium administration (arrow). D-F) An anterolateral bone infarction is shown in sagittal STIR, and axial T2-weighted images from Case 6. The sagittal STIR sequence (D) reveals a longitudinally extensive spinal cord lesion along with increased signal abnormalities in the T10 vertebral body (arrow). The axial T2-weighted image at the T10 level (E) demonstrates a large area of abnormal increased signal intensity affecting the left anterolateral aspect of the vertebra (arrows), as well as a thick-walled aortic atheroma with a lucid lipid core (arrowhead). Both axial images of the spinal cord in panels E and F highlight the selective signal abnormality in the anterior gray matter.

**Figure 4.**
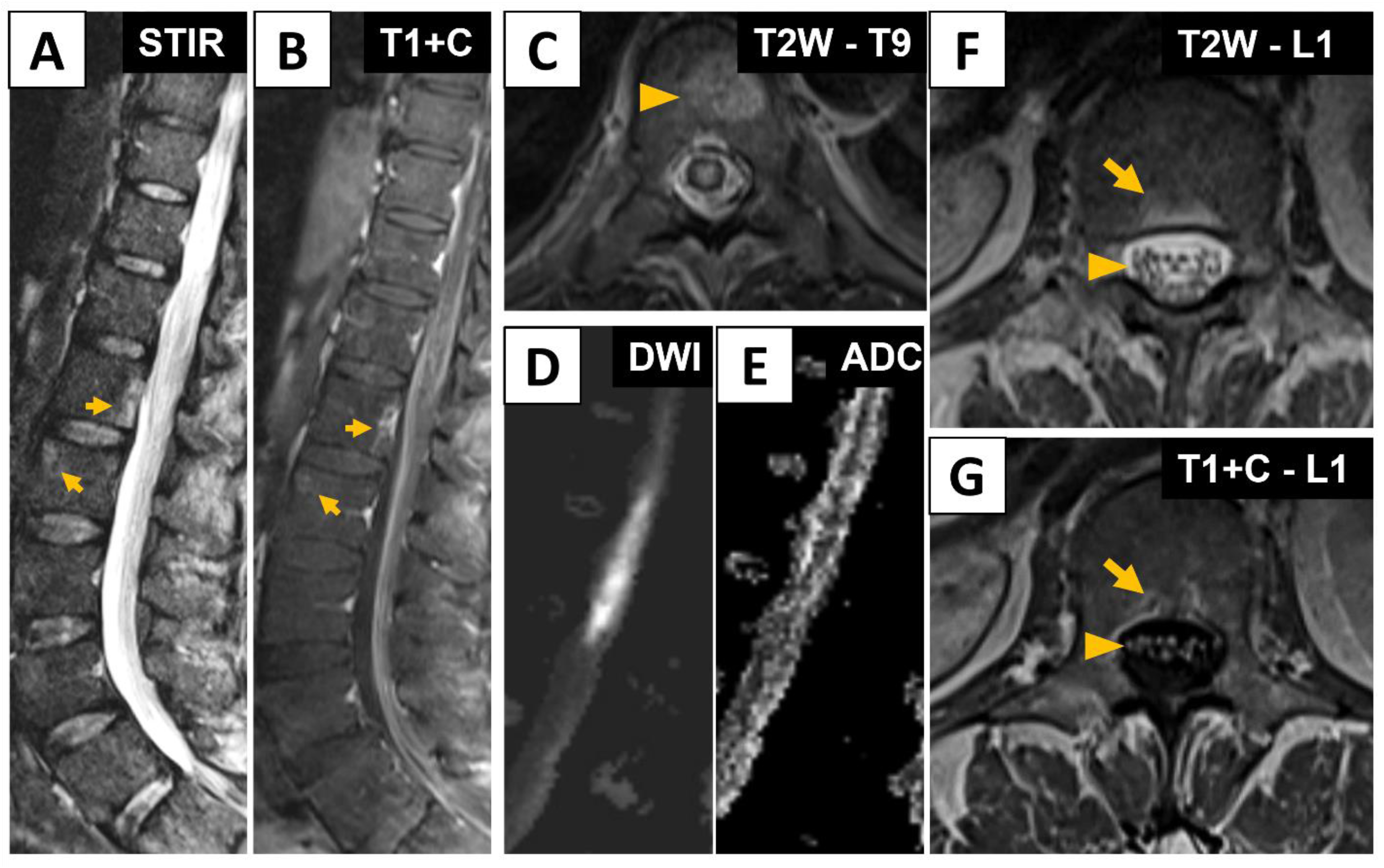
Posteriomedian and anterolateral vertebral bone infarctions in lumbar vertebrae. Vertebral bone infarctions in the lumbar spine, along with associated involvement of the spinal cord and cauda equina in Case 7. A) Sagittal STIR images reveal vertebral body infarctions located at L1 (posteromedian pattern) and L2 (anterolateral pattern), indicated by arrows. B) Corresponding Gadolinium-contrasted T1-weighted image shows matching areas of enhancement at these focal bone ischemic regions (arrows). C) A T2-weighted axial image at T9 vertebral body reveals a lesion with features suggestive of a hemangioma (arrowhead). D) Sagittal DWI illustrates focal areas of spinal cord ischemia in the lumbar enlargement (T11 to L1). E) An axial T2-weighted image at L1 shows the posteromedian bone infarct (arrow) and the cauda equina (arrowhead). F). A corresponding Gadolinium contrasted image demonstrates enhancement in both bone (arrow) and the anterior nerve roots (arrowhead).

**Figure 5.**
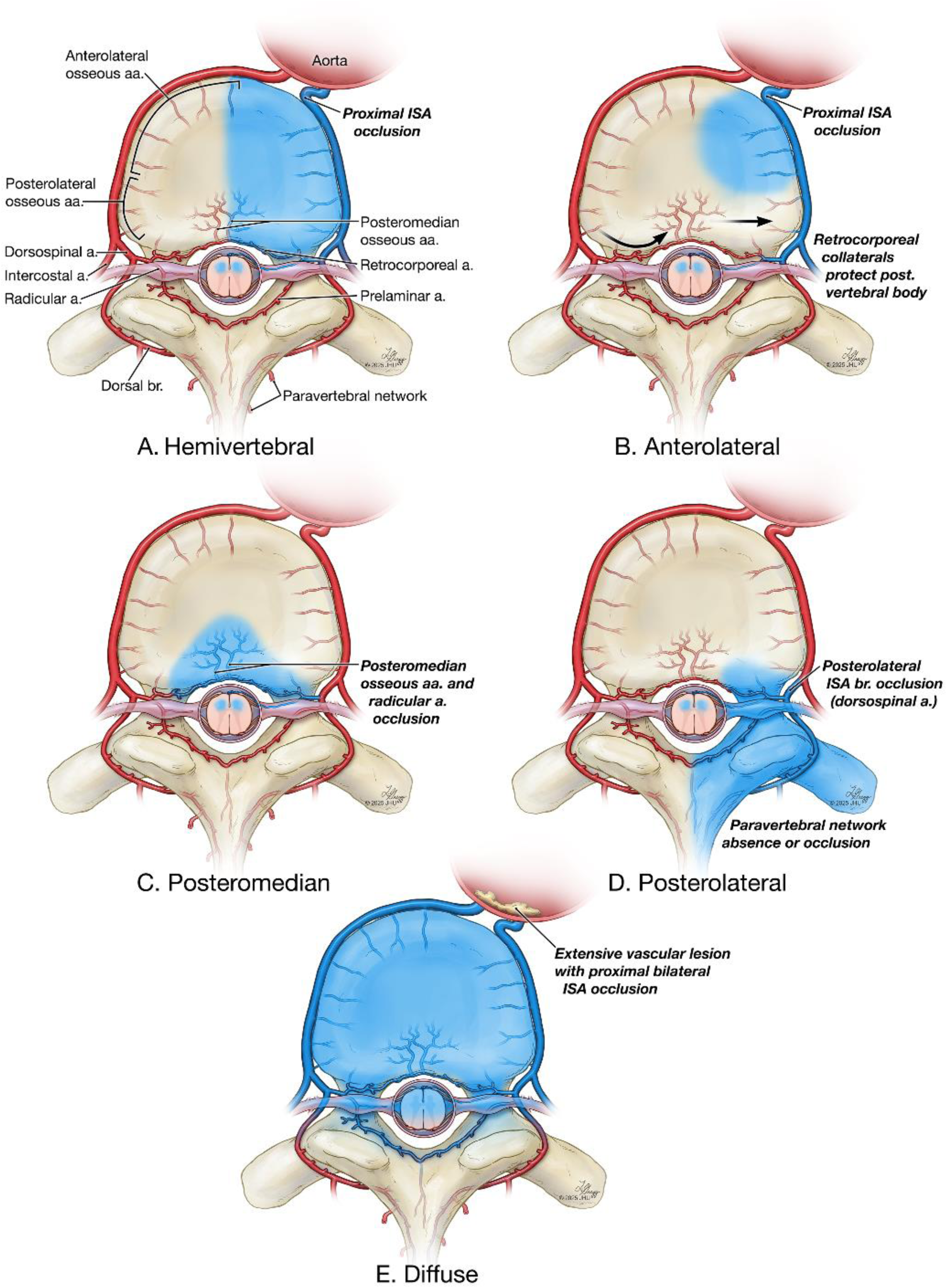
Patterns of Vertebral Bone Infarcts. Schematic illustration depicting vertebral bone infarction (VBI) topography as related to the arterial blood supply in the thoracolumbar spinal cord. **A.** Hemivertebral pattern of VBI associated with intersegmental artery (ISA) occlusion with inadequate collateral flow from the retrocorporeal artery and radicular branches. The posterior vertebra receives adequate flow from the paravertebral arterial network. **B.** Anterolateral pattern of VBI suggesting an injury to the ISA stem (i.e., ostial stenosis or diaphragmatic crus compression) resulting in compromised flow to the anterolateral group of osseous arteries. Collateral flow from the retrocorporeal artery and paravertebral arterial network protects the posterolateral vertebral body. **C.** Posteromedian pattern of VBI secondary to an occlusion or hypoperfusion to the medial branch of the ISA, affecting the radiculomedullary and retrocorporeal arteries. **D.** Posterolateral pattern of VBI with involvement of the pedicle, posterior arch, and spinous process, associated with occlusion or hypoperfusion of the posterolateral branches of the ISA coupled with an absent or compromised paravertebral arterial network. **E.** Diffuse pattern of VBI, associated with extensive vascular lesions affecting ISA ostia (e.g., atherosclerotic plaque). The posterior vertebra receives adequate flow from the paravertebral arterial network.

**Figure 6.**
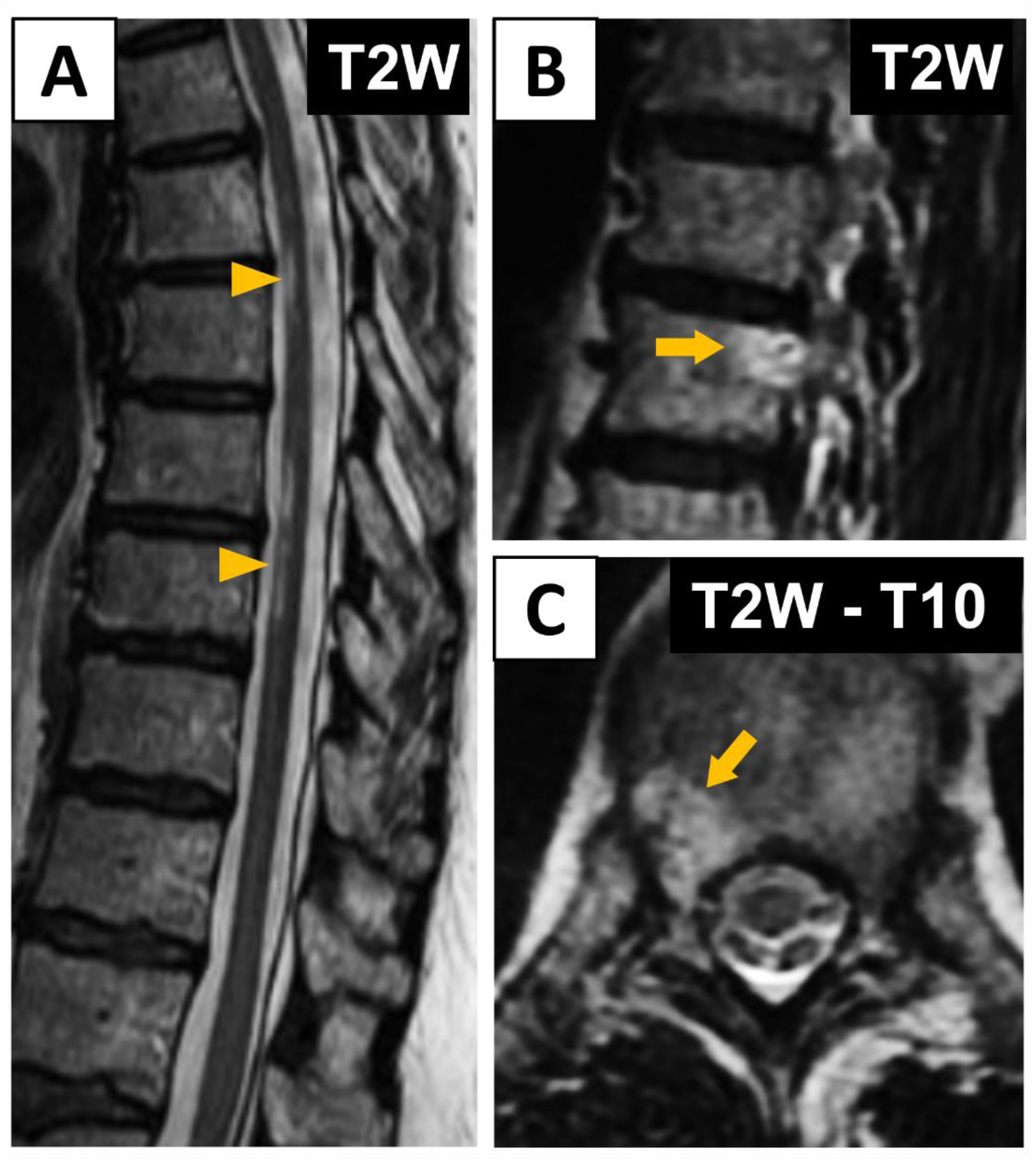
Spinal cord and vertebral bone infarcts with a posterolateral and pedicle bone infarction pattern. A) Sagittal T2-weighted image depicting patchy extensive spinal cord strokes (arrowheads) in the upper and middle thoracic spinal cord in Case 14. B) Sagittal T2-weighted image discloses posterolateral signal abnormality (arrow) at T10 consistent with BVI C) Axial T2-weighted image demonstrating a signal abnormality in the right posterolateral areas of the vertebra (arrow).

**Figure 7.**
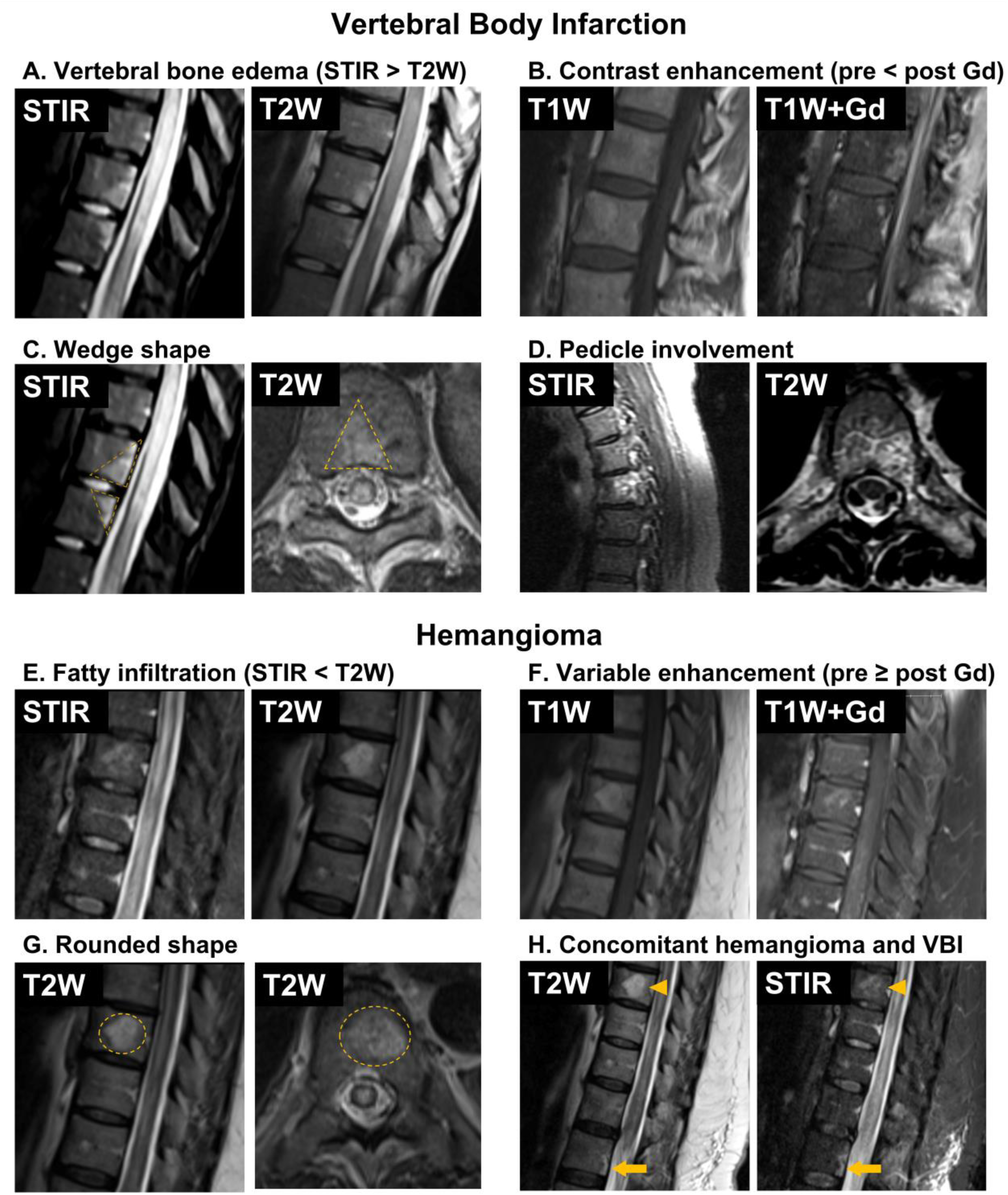
Key Imaging features of vertebral bone infarctions vs. vertebral bone hemangiomas. A-D) MRI features of vertebral bone infarction (VBI). A) Vertebral bone edema is identified as increased signal intensity, which is more clearly observed in STIR sequences compared to T2-weighted sequences. B) VBI is frequently visualized in Gadolinium-contrasted T1-weighted sequences. C) Characteristic wedge-shaped signal abnormalities are typical of VBI. D) In cases of posterior vertebral involvement, pedicle involvement can be observed (arrows). E) MRI features of vertebral bone hemangiomas. Fatty infiltration, which appears brighter on T2-weighted images than on STIR sequences, is a distinctive feature of bone hemangiomas. Hemangiomas display varying levels of Gadolinium enhancement, with vertebral bone hemangiomas typically appearing rounded on T2-weighted images. The signal abnormality rarely affects the surface of the vertebral body. F) Hemangioma and VBI may coexist. This image illustrates a hemangioma (arrowhead) and a VBI (arrow) in Case 7, demonstrating that both lesions can be present simultaneously.

## DISCUSSION

The main goal of this study was to assess the frequency of VBI in cases of spontaneous SCS, using a large cohort of patients diagnosed with this condition. Because SCS is often missed during the acute phase and misdiagnosed as “transverse myelitis” ^2^, there is a need for improved imaging techniques and clinical awareness of this acute myelopathic problem.

Identifying VBI could serve as an imaging biomarker for SCS, helping distinguish it from other myelopathies, such as myelitis, and prevent misdiagnosis. Our study examined the clinical and neuroimaging profiles of 15 patients with both SCS and VBI, representing 12% of the overall cohort of 126 patients with SCS, correlating MRI findings with relevant spinal vascular anatomy. A total of 35 vertebral body infarcts were identified in these patients. Most were initially misdiagnosed with inflammatory myelopathies, indicating that VBI was often overlooked or mistaken for spinal degenerative disease. This highlights the importance of considering VBI as a diagnostic marker for SCS, which could help avoid unnecessary immunosuppressive treatments and guide appropriate investigations and management plans. Although VBI was present in only 12% of SCS cases in this study, this frequency is slightly higher than previously reported, including a study describing 4 (7%) cases of VBI among 59 periprocedural SCS patients and 11 (9%) cases among 126 SCS patients. ^6^ However, it is less frequent than reported in smaller studies. ^4^ These frequencies are derived from retrospective studies that systematically evaluated VBI prevalence across diverse populations and criteria.

Our study findings have significant implications for both clinical practice and neuroimaging. Firstly, they emphasize the importance of a comprehensive clinical evaluation of patients presenting with hyperacute or acute myelopathic symptoms, particularly to recognize early stages of spinal cord ischemic injury and its potential association with vertebral bone ischemia. Secondly, our results highlight the value of detecting vertebral bone involvement, which may serve as a useful imaging biomarker for SCS. Lastly, the findings provide anatomical clues for localizing the site of vascular occlusions or underlying arterial pathology and understanding of the pathophysiology of some of the vascular mechanisms associated with SCS. It is not surprising that VBI is more common among patients with aortic atherosclerosis ^11^, as this condition affects the aortic origin of intersegmental arteries, directly affecting blood flow and restricting potential collateral sources. The extensive networks of paravertebral osseous and muscular arteries serve as significant protective factors, likely contributing in part to the relatively low occurrence of VBI among patients with SCS. The well-developed paravertebral networks contrast with the limited collateral supply to the spinal cord. ^12^ In this case series, which primarily involved patients referred from other centers with suspected inflammatory myelopathy, we observed a wide range in the time between presentation and the first MRI. Notably, VBI can be detected as early as 5 hours after symptom onset. This finding is consistent with previous studies that identified VBI within 5 to 12 hours of symptom onset. ^13,14^ In some cases, VBI may be the only abnormal finding on imaging for patients whose clinical presentation suggests spinal cord infarction. ^5^ Conversely, VBI can also be a delayed finding, becoming apparent several days after an initially normal MRI. In such scenarios, VBI serves as a confirmatory sign of SCS. ^7^

Identifying VBI on neuroimaging can be challenging, as its imaging characteristics often resemble those of degenerative changes, metastatic disease, or primary tumors, as well as frequent benign vascular abnormalities, such as hemangiomas, which may be present in vertebral bodies.^15^ Although not definitive, a combination of imaging findings can suggest the presence of VBI. As illustrated in our study and summarized in Figure 7, key MR imaging features are present in VBI. First, a notable finding was vertebral edema, best visualized on STIR images, an MRI sequence that suppresses fat and allows more precise differentiation between edema and normal bone marrow.^16^ Second, contrast enhancement after Gadolinium administration, which typically affects an area smaller than that documented by the STIR sequence, can aid in the diagnosis. Third, the presence of a wedge-shaped bone lesion, which is more indicative of ischemia than degenerative disease, ^9^ and further supports the MRI diagnosis of VBI. Finally, a signal-intensity abnormality extending into the pedicle and spinous process can also help differentiate VBI from focal vertebral body pathologies. While STIR and T2W MRI sequences provided valuable clues about vertebral bone edema or fatty infiltration for distinguishing VBI from hemangiomas, T1-weighted sequences were less reliable for detecting VBI due to inconsistent or absent signal changes, as shown previously ^9^ Although T1 hypointensity is typically associated with bone marrow edema, T1 hyperintensity, observed in two of the cases described, may suggest a hemorrhagic component. Importantly, MRI signal abnormalities consistent with the anatomical distribution of osseous arterial supply can help refine the diagnosis by identifying potential sites of arterial blood supply disruption, as illustrated in Figure 5.

In this study, VBI was most frequently observed at the thoracic and lumbar levels, below the upper limit of the cord lesion. Only one case showed a VBI above the cord lesion in the upper cervical region. This finding, previously noted in other studies, ^11^ can be attributed to two key anatomical features. First, the longitudinal supply area for each radiculomedullary artery in the cervical region is shorter than in the thoracolumbar region, where the artery of Adamkiewicz provides primary blood supply from approximately T6 and the conus medullaris. Second, during fetal development, the spinal cord and vertebral segments are aligned. However, as individuals mature into adulthood, a craniocaudal mismatch positions the conus medullaris around L1, leading to the formation of the cauda equina. Consequently, the radiculomedullary arteries and veins take an oblique course, following the lumbosacral nerve roots from the neural foramen to their corresponding spinal cord segments. The anatomical mismatch between the spinal cord and vertebral segments increases from the cervical to the lumbar regions. For example, the C6 cord segment aligns with the C5 vertebra, the T9 segment with T7 vertebra, and the L4 segment with T12 vertebra. This anatomical divergence explains why the levels of VBI and SCS are closer in cervical injuries and further apart in thoracolumbar injuries.

The common occurrence of VBI in two adjacent vertebrae within this cohort can be explained anatomically. In the anterior aspect of the vertebra, unilateral arterial trunks that supply two or more adjacent vertebrae can originate from a single ISA. Therefore, occlusion of a single ISA can result in ischemia affecting contiguous vertebrae, as illustrated in some of the cases described (Figure 5). In contrast, the posterior aspect of the vertebral body receives its blood supply from a dense retrocorporeal arterial network, which provides some protection against ischemia. However, this protective network becomes less effective with age, as the small arterial branches that cross the intervertebral space may be compromised by osteodiscal disease. Despite this, posterior VBI can still affect multiple vertebral levels,^5^ as seen in one of the cases described, where posteromedian involvement was observed at two contiguous levels (Figure 4).

The topography of VBI and the patterns of vertebral body involvement provide clues into the underlying physiopathology and site of arterial flow impairment, as illustrated in Figure 2. A proximal occlusion of the ISA is likely to result in a hemivertebral pattern of VBI, possibly due to insufficient collateral flow to the posterior vertebra or more severe ischemia. Infarction of the anterior region of the vertebral body suggests involvement of the anterolateral group of osseous arteries, indicating a possible occlusion of the ISA stem, such as ostial stenosis or compression by the diaphragmatic crus. In such instances, retrocorporeal collaterals typically may provide blood supply and protect the posterior vertebral body. Isolated posterior VBI is more likely to occur if there is injury to the medial branch of the ISA, which affects the radiculomedullary and retrocorporeal arteries (i.e., the posteromedian group of osseous arteries). A posterolateral infarct of the vertebral body, particularly if it is accompanied by pedicle involvement, suggests involvement of the posterolateral branches of the ISA. VBI that affects two or more non-adjacent vertebrae and various osseous territories indicates a more extensive vascular lesion, such as an aortic dissection ^17^, or significant central hypotension, particularly when associated with underlying aortic disease, as seen in two of the cases described. SCS without concurrent VBI typically occurs when arterial lesions are located downstream from the osseous supply, such as in cases of osteodiscal compression of the anterior spinal artery, or when collateral pathways sufficiently compensate for the compromised blood flow.

Individual variations in vascular anatomy and the robustness of collateral pathways explain the heterogeneity of osseous lesions. For example, a low lumbar lesion at the ostia of the ISA would be expected to cause bilateral VBI because bilateral ISA trunks are more common at this level. Similarly, unilateral VBI affecting adjacent vertebrae is anticipated to occur more frequently in the upper thoracic region. However, none of the lower thoracic or lumbar infarcts in this series involved bilateral anterolateral osseous territories. Extensive vascular lesions affecting the bilateral arterial supply, such as the vertebral arteries or ISAs, can result in a diffuse presentation of VBI, in which the posterior vertebrae still receive adequate blood flow due to a robust paravertebral arterial network. In the absence of this network, infarction may extend to the pedicle, posterior arch, spinous process, rib ^18^, or paraspinal muscles ^19^, as seen in some of the cases described. The cervical spine has a unique vascularization pattern, receiving blood from multiple sources, including vertebral, subclavian, and external carotid branches. This complexity means that, while cervical VBI remains an important diagnostic sign, arterial injury patterns are harder to determine than at the thoracolumbar level.

While our study has several strengths, including the evaluation of VBI frequency in a large cohort of patients with SCS and the use of a systematic approach to identify VBI using neuroradiological criteria, it also has limitations. First, it is a retrospective observational study rather than a prospective, systematic neuroimaging study of SCS evolution. This methodological approach limits our ability to draw definite conclusions about the natural history of VBI in the context of SCS. Second, since most patients with myelopathies typically undergo MRI of the cervical and thoracic spine, the absence of lumbar spine MRIs in some cases may have hindered the identification of VBIs at those levels. This is particularly significant because, in cases of SCS involving the lower thoracic and conus medullaris, arterial flow impairment may originate in the lumbar or pelvic blood supply. Third, the timing of image acquisition may have introduced variability in our findings and potentially prevented the detection of some VBIs.

Characteristic features of VBI usually manifest hours to days after ischemia onset, complicating the identification of VBI on MRIs performed too early or too late. Furthermore, Gadolinium enhancement may decrease, and the lesion may appear hyperintense on T1-weighted MRI due to hemorrhage or fat replacement. To address this issue, we reviewed all MRI studies obtained during the first month of symptom onset to increase the likelihood of detecting VBI, while also analyzing later MRI studies to detect older lesions.

## CONCLUSION

The co-occurrence of SCS and VBI is attributable to their shared arterial supply. This study highlights the significance of VBI as a critical MRI finding that supports the diagnosis of SCS in patients being evaluated for myelopathy, particularly in cases of hyperacute onset. It is essential to include VBI in the differential diagnosis of hyperintense vertebral lesions observed on T2-weighted sequences, particularly on STIR sequences. The MRI abnormalities associated with VBI typically exhibit patterns consistent with the underlying osseous blood supply. Increasing awareness of this association can facilitate accurate diagnosis and appropriate treatment, thereby minimizing the risk of misdiagnosis and unnecessary medical treatments.

## Data Availability

Data reported in this manuscript will be available upon request

## ACKNOWLEDGMENT

We want to thank the Bart McLean Fund for Neuroimmunology Research for its funding support. We are grateful to our patients and families who facilitated a comprehensive evaluation, and to our colleagues in the Department of Neurology, the Division of Neuroimmunology and Neurological Infections, and the Johns Hopkins Myelitis and Myelopathy Center for their contributions to the evaluation and follow-up of our patients.

